# Prominent Spatiotemporal Waves of COVID-19 Incidence in the United States: Implications for Causality, Forecasting, and Control

**DOI:** 10.1101/2021.06.29.21259726

**Authors:** Hawre Jalal, Kyueun Lee, Donald S. Burke

## Abstract

Better understanding of the spatiotemporal structure of the COVID-19 epidemic in the USA may help inform more effective prevention and control strategies. By analyzing daily COVID-19 case data in the United States, Mexico and Canada, we found four continental-scale epidemic wave patterns, including travelling waves, that spanned multiple state and even international boundaries. These major epidemic patterns co-varied strongly with continental-scale seasonal temperature change patterns. Geo-contiguous states shared similar timing and amplitude of epidemic wave patterns irrespective of similarities or differences in state government political party affiliations. These analyses provide evidence that seasonal factors, probably weather changes, have exerted major effects on local COVID-19 incidence rates. Seasonal wave patterns observed during the first year of the epidemic may become repeated in the subsequent years.

**One Sentence Summary:** The COVID-19 epidemic in the United States has consisted of four continental-scale spatiotemporal waves of case incidence that have spanned multiple states and even international boundaries.

---

Since the first case of COVID-19 was reported in Wuhan, China in December 2019, an ongoing global pandemic of COVID-19 has led to 180 million cases and 3.9 million deaths (*1*). In the United States, the national epidemic incidence has waxed and waned four times, with varying intensity in different regions of the country (*2*). Possible causes for changing case rates include changes in social distancing, mask wearing, and other epidemic control polices; emergence of viral variants; changes in the weather; introduction of vaccines; and other factors.

Spatiotemporal analysis of COVID-19 can provide improved understanding of patterns and drivers of the COVID-19 epidemic in a given geographic region. By comparing epidemic patterns across the country, it is possible to identify clusters of locations that share similar patterns. For example, Kang et al showed the spread of COVID-19 from the Hubei province where the first outbreak occurred in China to neighboring provinces (*3*). Another study in Brazil traced the trajectories of the geographic center of the epidemic over time to understand the spread of COVID-19 in the country (*4*). For the US, Zhu D et al used a network modeling to examine the spatial shifting patterns of COVID-19 from February to August in 2020 (*5*). As more than a full year of data are available now, the spatiotemporal patterns of COVID-19 can reveal the association between geographical properties and the spatiotemporal patterns of COVID-19. Furthermore, spatiotemporal pattern can be better understood, if the pattern is compared with the neighboring countries such as Canada and Mexico in case of the USA.

## Results

The case and mortality rate curves in the US, Mexico and Canada reveal four dominant waves over the period from 03/01/2020 to 05/03/2021 (**Fig. 1**). These waves are also discernable in most states and provinces in these countries (Supplementary **Fig. S1** and **Fig. S2**). The first wave of epidemic began in late March 2020, the second wave around mid-June 2020, the third wave started in the beginning of September 2020 and peaked with the largest amplitude compared to other waves during the coldest months of a year in December 2020 and January 2021. The fourth wave occurred in spring in late March and early April in 2021, around the same timing of the first wave in 2020. Since the timing and amplitude of mortality rate waves were highly correlated with those of case rate wave (Supplementary **Fig. S3**), in this analysis we primarily used case rates to better capture regions with smaller populations.

**Fig. 1.**
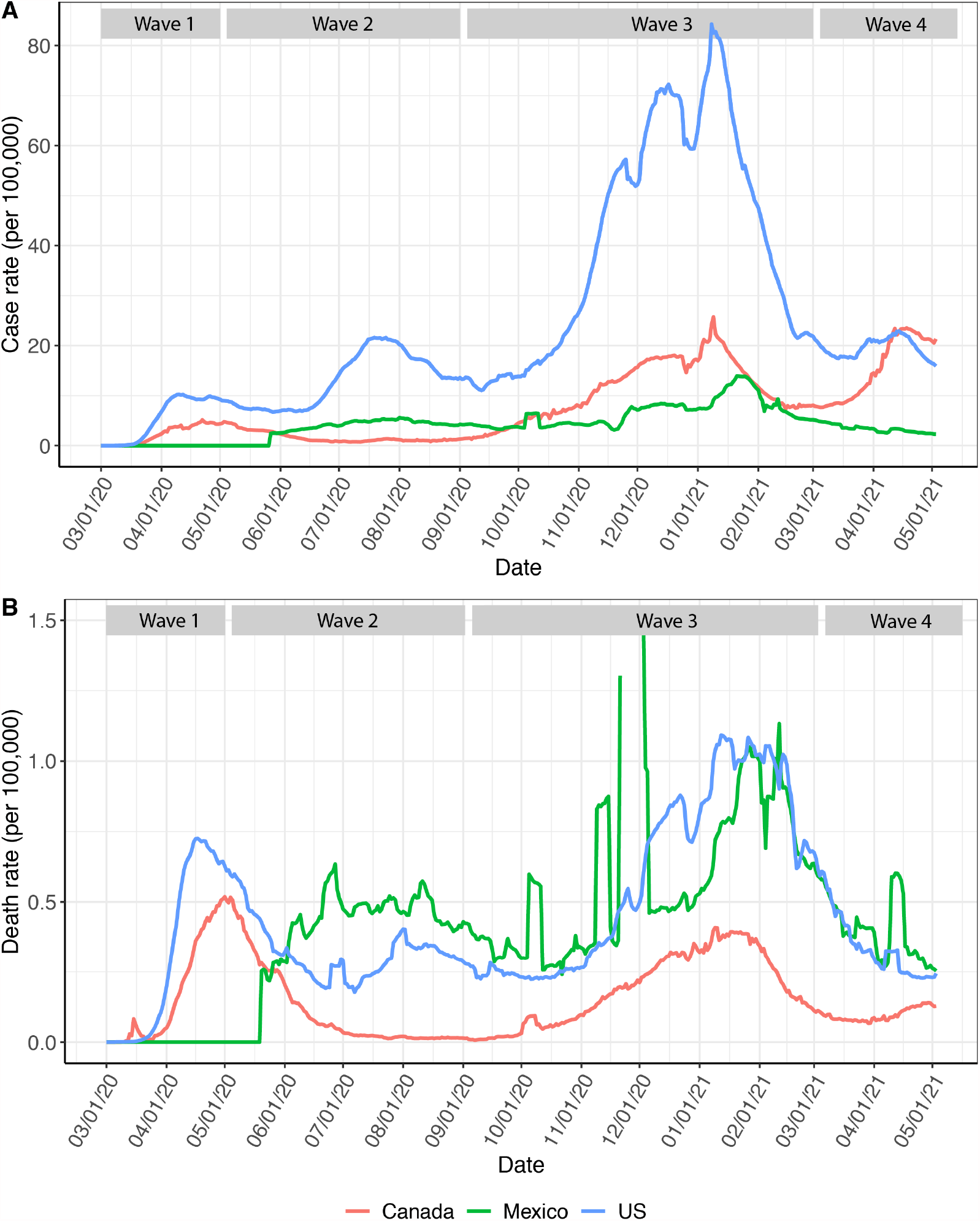
The four waves of COVID-19 cases and mortality for US, Mexico, and Canada. Panel A shows case rates, and Panel B shows Mortality rates are per 100K. Blue, green, and red line indicates 7-day average of case or mortality rate per 100,000 in the US, Mexico, and Canada, respectively. The discontinuity in Mexico’s mortality curve is because the y-axis is limited to less than 1.5 per 100,000 which suppresses the spike in reported deaths around 12/01/2020.

**Fig. 2, Movie 1**, and **Movie 2** were constructed to allow visualization of the spatiotemporal patterns of Covid-19 in the USA and how the patterns in the USA related to those in Canada and Mexico. To describe this analytic approach, we purposely borrow the term “synoptic” from “synoptic meteorology” to emphasize the scale of the epidemic analysis. Synoptic meteorology is routinely used to describe the analysis of weather patterns at continental level at scales of 1000 kilometers across or more.

**Fig. 2.**
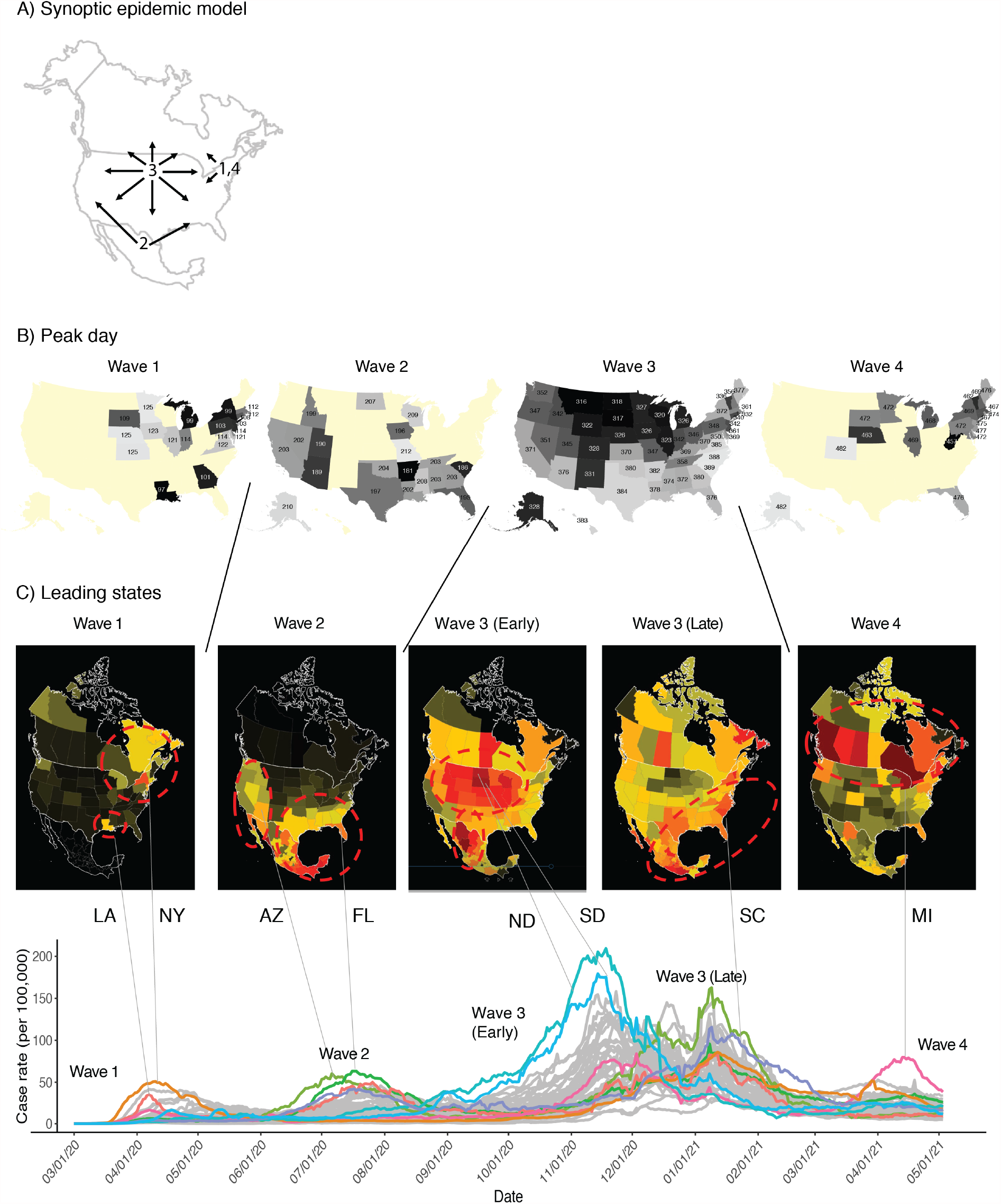
Synoptic Epidemic Model of Covid-19 in the USA: (A) shows a illustrates the overall synoptic model with numbers indicated waves 1 through 4, and arrows depicting the general chronological order of states involved in each wave. (B) shows the peak day for each wave by state from the Gaussian function fit (Supplementary **Fig. S7**) where darker colors represent earlier states in that wave, and (C) shows the states leading each wave.

Wave 1 (1 March to 31 May 2020 = days 61 to 152) started in New England and Eastern Canada, with subsequent scattered intensity in the northern half of North America. Wave 2 (1 June 2020 to 30 August 2020 = days 153 to 244) displayed a travelling wave pattern. It first appeared in Mexico, crested northward to the southern and western USA states, then reversing in intensity surge and ebbing back southward. Wave 2 first appeared in the USA in Arizona, then moved northward along the West Coast and eastward into Florida and other Gulf Coast states. Wave 3 (1 September 2020 to 28 February 2021 = days 245 to 425) emerged as a travelling wave from the Dakotas, flowing outward in a radial pattern to essentially all North America. The ebb of Wave 3 followed along the same general spatial pattern as its flow, again starting in the Dakota region, and ebbing outward in a radial pattern across the whole continent. Wave 4 (1 March 2021 to at least 4 May 2021 = days 426 to 489+) emerged from the tail end of Wave 3 in New England and rose quickly across all of Canada and some northern USA states.

To reveal a more detailed synoptic mapping of COVID-19 epidemic in the USA, we created a second movie (**Movie 2**) of county-level case rate data. This movie displays the z-scores from a hot-spot analysis using a modified Getis-Ord Gi* statistics (*6*). The red colors indicate counties and their neighbors being above the national average and the blue colors represent counties that are below this average. In addition, the modified Gi* allows the hotspot analysis to be conducted through time as well as spatially (*7*). For example, close observation of Wave 3 shows the gradual county by county movement of the epidemic from west to east from Iowa through Illinois, Indiana, Ohio, and Pennsylvania to New York and New England over the period 15 October through 15 December 2020.

Next, we quantitated the wave movement patterns that were apparent visually in the movies (**Fig. 2**). This figure presents a summary of the synoptic mapping of the geospatial patterns for all the waves. By representing the case time series for each state as log-linear model with multiple change points, we were able to estimate the peak day of all four waves in each state (Supplementary **Fig. S4**). Estimation of the differences in peak days between pairs of states serves as a measure of the timing, in days, of the state-to-state movement of a wave between those states. In addition, comparing the amplitude of the estimated peaks for each wave provides a measure of case severity. Because not all states were involved in all four waves, for waves 1,2, and 4, we focused on the timing patterns among only the 20 most affected states (the twenty with the highest case rates) for that wave.

In wave 1, Louisiana shows an early peak (day 97). New York and Michigan peaks (day 99) were followed by other states contiguous to New York (days 103-108). Then, Georgia peaked (day 101) and the north central states peaked (days 109-125). Wave 1 lasted about one month.

In wave 2, Utah and Arizona peaked early (days 189-190). Although, the figure shows Arkansas and South Carolina peaking earlier (days 181 and 186, respectively), examining the change point fits carefully (**Fig. S4**), reveals that the case rate rose quickly in these two states, but the curves actually peaked after day 200 in wave 2. Then, the southern shoreline states (FL, TX and LA) peaked (days 194-202), followed by other shoreline states and the adjacent southern inland states (GA, AL, TN, OK and MS, days 202-207). Collectively the epidemic in these highly affected southeastern states in wave 2 formed a northward travelling wave of peak intensity that lasted about a month. In the far west, Idaho, California, and Nevada (days 199-203) also showed increased wave 2 intensity.

Wave 3 affected all the USA states. North Dakota, South Dakota, and Montana peaked first on days 316-318. The wave 3 epidemic peaked next in nearby north central states (WI, WY, IL, IA, MI, NE and MN; days 320-326), and proceeded to move radially outward through successive tiers of states. For example, the wave 3 peak epidemic day of states along north to south transect through states in the center of the country (MT, WY, CO and NM) had successively later peak days of 316, 322, 328 and 331. The crest of the travelling wave was followed by an ebb wave, again from north to south.

We only had partial data on Wave 4 at the time of writing this manuscript. Thus, the peak days for wave 4 are less stable (**Fig. S4**). Wave 4 overlapped with the final stages of wave 3. As in wave 1, wave 4 first peaked in the northeastern states followed by north central states, then, other states scattered across the country.

To further characterize the case rate dynamics both nationally and internationally, we compare the proximity between each state pair in both case rate trend similarity and geographical proximity in the USA, Canada and Mexico. We show that states that are geographically closer to each other are more likely to have similar patterns of COVID-19 epidemics, compared to those located farther apart (**Fig. 3**). Correlation between COVID-19 case rate distance and geographical distance for all pairs of states in US, Mexico, and Canada was 0.47 (p<0.001) (supplementary **Fig. S5**). The level of this correlation varied by the index state. For example, **Fig. 3** shows the correlation between case-rate distance and geospatial distance where the index states were South Dakota in the US, Veracruz in Mexico, and Manitoba in Canada. These states present strong spatial relationships of case time series and geographical distance to the other states. We also show the case-rate and geographical distance for California (USA) where the position in case series space does not respect the position in the geographical space. It is notable that Veracruz (Mexico) was closer to other Mexican states in case rate distance, followed by southern USA border states such as Louisiana, Texas, Florida. The pattern was similar for Manitoba in Canada which is closer to the northern USA border states such as Minnesota, South Dakota, and North Dakota compared to other states in the USA and Mexico.

**Fig. 3.**
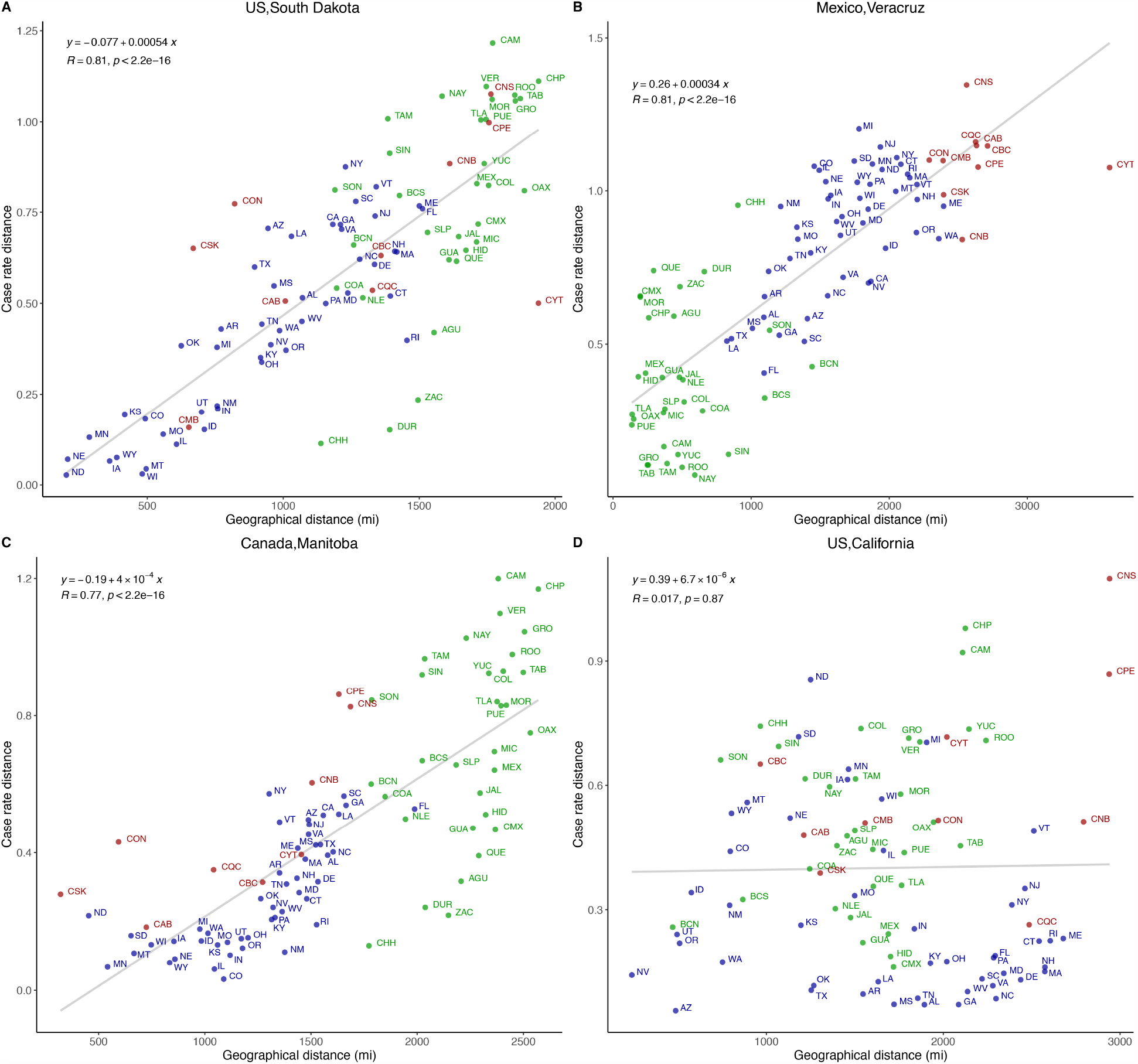
Correlation of case time series distance to geographical distance in four index states: (A) South Dakota, US; (B) Veracruz, Mexico; (C) Manitoba, Canada; and (D) California, US. The linear fit, correlation coefficient (R) and the p-value of the fit are shown for each panel. [A description for the state and province abbreviations used are presented in the Supplementary **Table S2**].

Next, we projected the similarity in case space into 2D and 3D space which revealed a pattern preserving major geographical relationships among states in the three countries (**Fig. 4** and **Movie 3**). The USA border states in the north and south were contiguous to the Canadian and Mexican states, respectively (**Fig. 4A**). Mexican states formed their own cluster in case distance mapping space which overlapped with the southern states USA states such as Florida, Texas, Arizona, Louisiana, and South Carolina. Chihuahua was apart from the major Mexican cluster, and it showed proximity in case distance space to northern Midwest states such as North Dakota, South Dakota, and Iowa. Canadian states spread out in the case distance space and most states bordered with the northern states of USA such as Minnesota, New York, New Jersey, Vermont, Massachusetts, and Maine. The case distance projections for the USA states were similar to the geographical map of the United States, except for the pacific states (CA. OR, WA) (**Fig. 4B**). States that are congruent in geographical space tended to have similar patterns of COVID-19 incidence rates in the US. Northeast USA states (NJ, NY, ME, MA, NH, MD, VT) and located in upper right corner of the case distance map, Southeast and Southwest states (TX, SC, GA, FL) clustered in the south, and Midwest states (ND, SD, WI, IA, IL) and Rocky Mountain states (CO, UT, WY, NV, ID, MO) are in northwest and mid-west in the space of case distance. However, certain groupings of states were displaced from their expected locations in case time series space. For this analysis, we used the map of the ten Regions of the USA Department of Health and Human Services. HHS Region 2 (NY and NJ) was modestly displaced northward; HHS Region 9 (CA, AZ, and NV) was displaced substantially eastward to cluster with the southeastern states; and HHS Region 10 (ID, OR, WA) was also displaced eastward to cluster with the mid-western states. Because HHS Region 9 and Region 10 are both on the West Coast, their combined displacements to the east in the case time series map can be approximated by “folding” the geo-map along the axis of the Rocky Mountains.

**Fig. 4.**
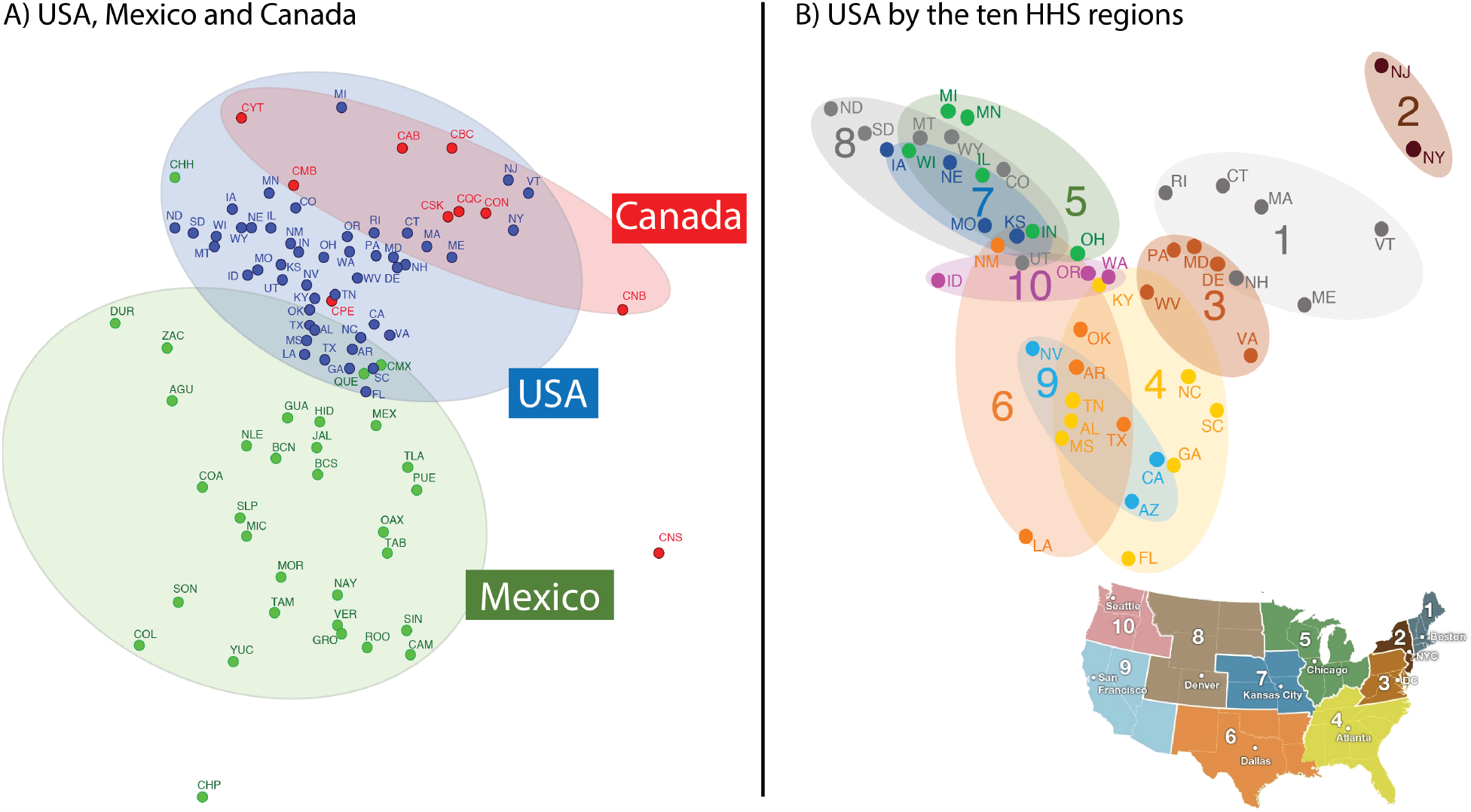
Two-Dimensional projection of all US, Mexican, and Canadian states in the space of epidemic case distance. In (A), states and provinces in all three countries are mapped and the shaded region groups the states based on the country. In (B), states in the USA are mapped in the space of case rate distance. We grouped the states based on the HHS regions and compared the position in the space of case distance with the geographical location. [A description for the abbreviations used are presented in the Supplementary **Table S2**.]

Given that neighboring states predictably have very similar epidemic *trajectories*, we hypothesized that neighboring states with different policy approaches might nonetheless have dissimilar epidemic *amplitudes*. Therefore, for every state, we measured the mean difference in wave amplitude between that given state and all of its neighboring states. Next, we identified neighboring state pairs where political control of the state was the same, or different, and compared the differences in epidemic amplitude between those pairs where political control was the same or different. Neighboring state pairings with Democratic/Democratic, Republican/Republican, and Split/Split political control were considered to have similar political control, whereas all other pairings were considered to have dissimilar political control. The average difference in peak case rates between dissimilar and similar pairs were 2.38 (p-value = 0.025, not corrected for multiple observations), −0.553 (p-value = 0.39), −1.86 (p-value = 0.72) and 0.677 (p-value = 0.63) for waves 1,2,3 and 4, respectively. Thus, we were unable to detect statistically significant systematic differences in epidemic intensity among geo-contiguous state pairs with politically similar versus politically dissimilar party leadership except for wave 1 which was early in the epidemic. Regardless of political leadership, geo-contiguous states had similar timings and intensities of their COVID-19 epidemic waves.

Given the apparent association of Wave 3 with the onset of winter, we explored the relationship between wave 3’s peak incidence day and the day when each state’s temperature reaches 15°C, 10°C, 5°C and 0°C (**Fig. 5**). The x-axis of this figure is the first day when the 7-day moving average of daily temperature reaches the specified threshold within +/-1°C. To represent temperature in populated areas, we weighted the average daily temperature in each squared kilometer by the proportion of the state’s population residing in that area. The y-axis represents the peak incidence day for wave 3 as measured by the change point analysis discussed above. The 95% credible intervals are also shown is light blue vertical lines. The diagonal line indicates that case incidences peak when temperature reaches the threshold. Most of the states fall above the line indicating that the day of reaching the specific temperature threshold occurs prior to the wave 3 case peak inflection. The correlation coefficients were 0.62, 0.71, 0.77, and 0.73 for temperature thresholds of 15°C, 10°C, 5°C and 0°C, respectively. Supplementary **Fig. S6** shows the average temperature trend for each state and the selected day in which the 7-day moving average temperature trend reaches 10°C as an example to illustrate the trends.

**Fig. 5.**
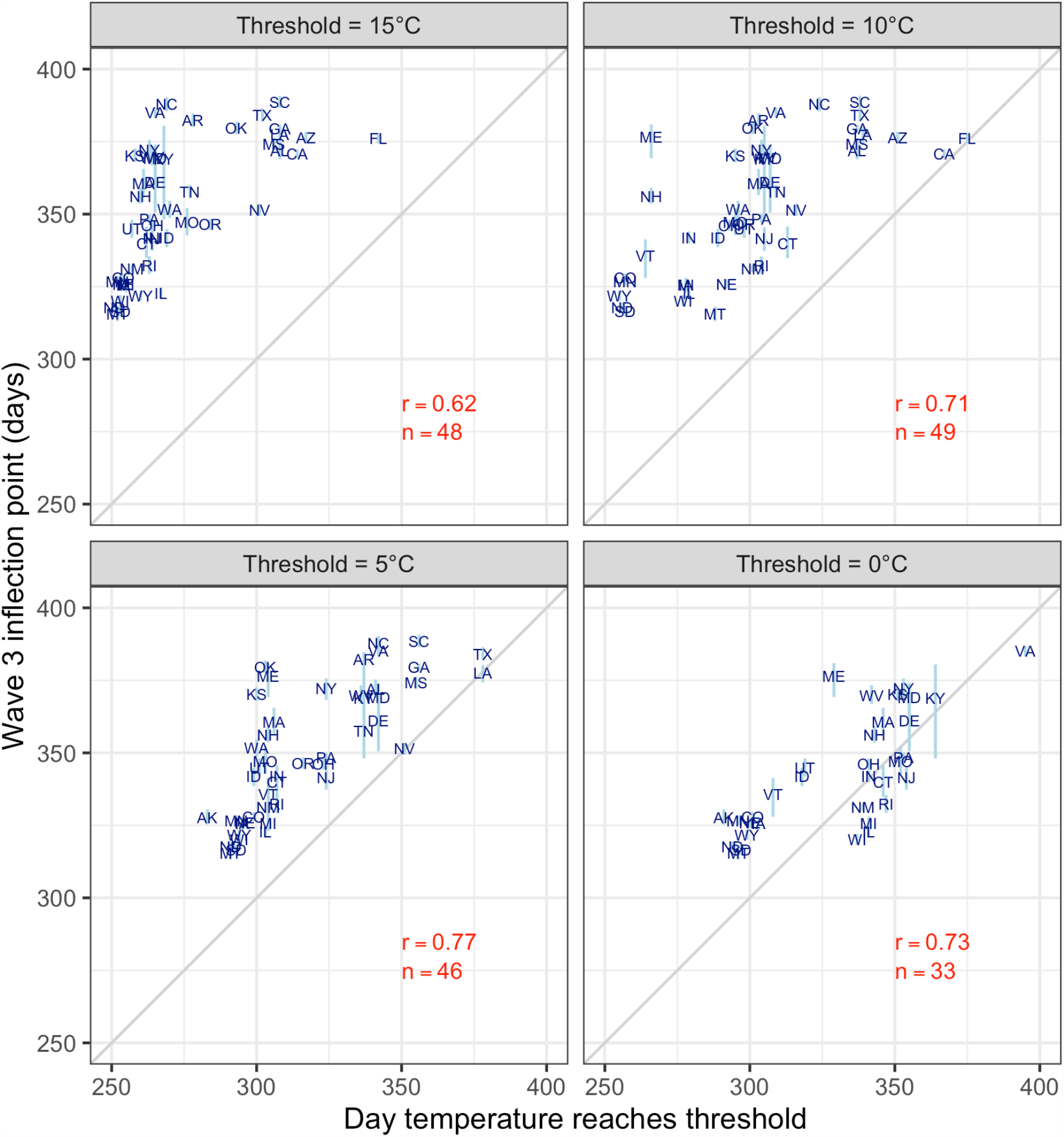
Association between average temperature and wave 3 inflection point. Each plot shows a specific temperature threshold. The x-axis displays when the 7-day moving average temperature in each state reaches this threshold within +/-1°C, and the y-axis is wave 3’s peak incidence day estimated from the change point analysis. The 2-letter abbreviations represent the USA states. The light blue vertical lines are the 95% credible intervals for each state’s inflection point. r is the correlation between temperature threshold day and the wave 3 inflection point, and n is the number of states that reach the threshold excluding Hawaii.

## Discussion

We used daily COVID-19 case data to visualize, detect, and analyze spatiotemporal patterns of the epidemic at a continental scale in North America. This “synoptic epidemic mapping” approach allowed us to detect considerable spatiotemporal structure of the four COVID-19 epidemic waves. By creating a full continental map of the normalized daily local intensity scores for all states and provinces for every day, and by displaying these 489 daily maps (1 January 2020 = day 1) in sequence, we created movies that revealed continent-wide changes in intensity at the state-level for North America (**Movie 1**) and at the county-level for the USA (**Movie 2**).

Both movies reveal the dynamic nature of the waves across state and national boundaries. Overall, wave 1 was the smallest and least structured of the four waves, starting in New England and Eastern Canada. Especially for epidemic movements within wave 1, it may be difficult to disentangle the effects of new introductions versus other factors more associated with increased transmission of an already established local epidemic. It has been speculated that New York and New Jersey were the first states to be severely affected because of their high population density and relative proximity to Europe through air travel (*8*). Note that early in wave 1, a brief ectopic focus of local intensity appeared in Louisiana and then neighboring Mississippi, possibly related to Mardi Gras on 25 February 2020. Wave 2 was more structured starting from the southern US states and traveling northward. Interestingly, the COVID-19 normalized local intensity appeared to increase sharply in several Mexican states in the days preceding its start in the USA. Wave 3 displayed the strongest spatial pattern of the four waves starting in the Dakotas, radially spreading to the entire North American continent, and then receding in similar fashion. Wave 4 emerged from tail end of Wave 3 in New England and rose quickly across all of Canada and some northern USA states. Given that the COVID-19 epidemic emerged in the USA only slightly more than one year ago, there is insufficient time series data to determine if the wave pattern observed during the first year will be repeated in future years. It is interesting nonetheless to note that the North American wave 4, one year later, appears to roughly spatially congruent with epidemic wave 1 along the USA / Canada border.

Calculating the pairwise differences in the daily reported COVID-19 time series cases for all states (and provinces) in the USA, Mexico and Canada allowed us to create and visualize maps of the distances among these regions in COVID-19 case time series space and compare those maps to conventional geo-maps. The state-to-state distance relationships in the COVID-19 case time series map were broadly congruent with their distance relationships in geo-space, even across international boundaries and datasets. Geo-contiguous state pairs were far more likely to show similar case time series patterns than geo-distant state pairs.

We performed one preliminary analysis of the effect of political party control of state legislatures and governorships on the amplitude of the COVID-19 epidemic. In the USA, state level epidemic control policies are strongly associated with the political party affiliation of the state government leadership. For example, Democratic-led states typically have more stringent policies for mask wearing and social distancing compared to Republican-led states (*9*). Our analysis effectively asked whether “red state / blue state” neighbor pairings were more likely to have dissimilar epidemic amplitudes than “red state / red state” or “blue state / blue state” pairings. For waves 2, 3, and 4, we could not find any association between neighbors’ state political differences and neighbors’ state epidemic amplitudes. Only for wave 1, early in the epidemic, we found a borderline significant association (uncorrected for multiple observations) of political difference and epidemic intensity difference. This preliminary analysis does not exclude the possibility that national and/or state policies could have lowered epidemic amplitudes across all pairings. We believe that this approach, of choosing neighboring controls, will prove useful in future COVID-19 policy analyses.

Our findings suggests that temporal variations in the epidemic intensity in a particular USA state are subject to forces acting well beyond state or national borders. Indeed, epidemic intensity at any given point in time can co-vary with the epidemic intensity in states a thousand miles away or more. This is especially true of states at the same latitude. For example, the case time series patterns for California and Arizona are very similar (r > .9) to those of Mississippi, Alabama, and Georgia. These wide-acting forces span international borders as well as state borders. The wave patterns also cross boundaries between red states and blue states.

The striking geospatial structure of the COVID-19 epidemic in the USA, dominated by shifting north south latitudinal gradients, strongly suggests a role for seasonal weather-related factors as major drivers of the epidemic. Our preliminary finding revealing a strong relationship between air temperature and peak case incidence rates in wave 3 (correlation > 75%) supports such role. There has been agreement on the significant role of temperature in COVID-19 transmission (*10*). Like other respiratory viruses, COVID-19 transmission rate is affected by environmental factors such as temperature and humidity. On the other hand, there are mixed observations on the relationship between relative or absolute humidity and COVID-19 transmission (*10*). Changes in both viral stability and in human behaviors have been associated with changes in environmental factors (*11, 12*). Environmental conditions determine how long the virus persists in aerosols and on surface (*13*). Increases in indoor human to human contacts during the cold weather can increase viral transmission between infected and susceptible individuals (*12*).

Although, latitude and altitude are known to be closely correlated with mean temperatures, the quantitative relationship between COVID-19 epidemic intensity and temperature is not a simple one. While wave 3 behaved as expected, with higher epidemic intensity emerging in the north and tracking southward as the winter season and cooler temperatures arrived, the earlier summertime wave 2 behaved exactly the opposite. Paradoxically, increased epidemic intensity in wave 2 first emerged in states in southern latitudes and with low altitudes, in the hottest states in the hottest time of the year. Studies identified the range of 5-15°C as a significant temperature zone for high incidence of COVID-19 cases (*14-16*). Outside this temperature range, COVID-19’s temperature response becomes unclear (*14, 16, 17*). Wave 2 also appears to have started in low altitude states in Mexico concurrently with the southern USA states. It appears likely that different seasonal-associated mechanisms may account for wave 2 and wave 3. This topic warrants further study.

The causes of transitory waxing or waning of COVID-19 case incidence rates have been the subject of prior analyses. Some studies support the effect of social-distancing measures such as closing schools and stay-at-home orders and other non-pharmaceutical interventions such as mask wearing on reducing the growth rate of infections (*18-22*). No doubt these measures among other factors can affect the state-by-state intensity patterns too, including travel, population density and demographics, viral variants, vaccine coverage, and other important factors. A proper causal analysis for the COVID-19 waves should include all these factors, and their interactions. Our hope is that this spatiotemporal framework will contribute to such definitive analyses.

Whatever the exact causal mechanism, it seems highly likely that wave 3 will occur again in the fall of 2021, given its strong correlation with temperature and seasonal pattern in 2020. It is also very possible that wave 2, given its strong continent-spanning spatiotemporal pattern, will occur again this summer in 2021. However, uncertainty about the possible mechanisms relating southern latitudes and low altitudes to the wave 2 surge lowers this expectation somewhat. Increased accumulation of immune persons in the population over the past year, through prior infections and through vaccination, will almost certainly dampen future epidemic waves in North America. However, as long as the population remains at immunity levels that are lower than those required for herd immunity, the seasonal forces that drove wave 2 and wave 3 could result in measurable epidemic surges at the same times and same regions in 2021 as occurred in 2020. Similar spatiotemporal analyses of other large scale COVID-19 data sets could be done around the world, linking the local epidemic patterns to local weather patterns. Results of such comparable international analyses could extend and/or support possible new epidemic forecasting methods based on weather.

This analysis suggests that the pattern of waxing and waning is largely driven by external factors. However, the overall amplitude (intensity) of the epidemic can nonetheless vary across states with identical time series patterns. It is not yet possible to quantitate exactly what proportion of the temporal variance in epidemic intensity in each state can be attributed to extra-versus intra-state factors. From these observations, we conclude that analyses to evaluate the effectiveness of state control policies should not be based solely on temporal changes of time series case reports within each state, because the pattern of waxing and waning temporal changes in that state is probably largely the result of forces that are external to that state. We propose that where possible, policy analyses should be done by comparing the pattern and amplitude of the case time series in a state to the pattern and amplitude of the case time series in neighboring states.

## Data Availability

All data used in this analysis is publicly available from the identified sources. The code used is available upon request from the authors.

## Funding

This research was supported by the Public Health Dynamics Laboratory (PHDL) at the University of Pittsburgh Graduate School of Public Health.

## Author contributions

Conceptualization: HJ, DSB

Methodology: HJ, KL

Investigation: HJ, KL

Visualization: HJ, KL

Funding acquisition: HJ, DSB

Project administration: HJ

Supervision: DSB

Writing – original draft: DSB, HJ, KL

Writing – review & editing: DSB, HJ, KL

## Competing interests

Authors declare that they have no competing interests.

## Supplementary Materials

Materials and Methods

Supplementary Text

Figs. S1 to S9

Tables S1 to S2

References (*7, 24-27*)

## Movie captions

**Movie 1**. Animation of normalized local intensity of COVID-19 cases in the US, Canada and Mexico. The color is scaled between 0 and 1 within state. Zero indicating no cases in a state and 1 indicates the maximum number of cases for that state during the observation period.

**Movie 2**. Animation of geospatial hotspot of COVID-19 case rates. The Gi* statistics are standardized using pooled statistics over time. The various shades of red and blue indicate pooled standard deviations above and below the pooled mean, respectively, as shown in the legend.

**Movie 3**. Animation of three-dimensional plot of all US, Canadian, and Mexican states in the space of case distance.

## Materials and Methods

### Data sources

This analysis uses COVID-19 case rates for Canada, US and Mexico from the start of the pandemic until May 3^rd^ 2021, the latest date available at the time of writing this manuscript. We obtained daily cases and deaths data by state from data compiled by The New York Times and John Hopkins University (*1*). We used 2020 population for all US and Mexico states and Canadian provinces. All rates are calculated per 100,000 people. We excluded Hawaii and Alaska for analyses that involved studying the contiguous US.

### Case distance vs. geographical distance

In order to examine whether geographically close states tend to have similar patterns (rise and wane) of COVID-19, we first estimated correlation between geographical distance and case distance in every pair of states in the US, Mexico, and Canada except for Hawaii and Alaska. In each pair of states, we calculated the Pearson’s correlation of the 7-day average of case rate from March 1, 2020 to May 3, 2021. The case rate distance was defined to be subtracting the absolute value of correlation from 1. Hence, the case rate distance will capture similarity and difference in patterns of COVID-19 epidemics in terms of the timings of the 4 waves and not the severity. Geographical distance was calculated based on the distance between the centroid of two states.

We also studied whether the network mapping of case distance can restore the geographical network structure of the states. We mapped states (vertices or nodes) in the 3-dimensional space based on their case distance (edge). We used Kamada-Kawai layout algorithm (*2*) to optimize the layout of network in a 3-dimensional space given the distance in case rates.

### Normalized daily local intensity score

To further reveal the geospatial patterns of COVID-19, we normalized daily local intensity score by rescaling the number of cases in each state relative to the maximum number of cases observed in that state over the study period. We produced an animation of these rescaled patterns to study the dynamics of changing local intensity in a state. To reduce the temporal noise, we used Locally Weighted Smoothing (LOESS). In the county-level analysis, we used the Getis-Ord Gi* statistic to show geospatial clustering of hot (high) and cold (low) spots of case rates. The Gi* statistic identifies these hot and cold spots on the basis of contiguous counties. The Gi* statistic is essentially a Z-score standardized by a mean and standard deviation of case rates in all the counties. Typically, the Gi* statistic can display geospatial information on one dimension such as mortality rates. To add additional dimensions and compare case rates over time, we re-standardized the Gi* statistics by using the pooled mean and standard deviations of the Gi* statistics over time (*3*). This re-standardization allowed us to produce a set of comparable maps over time in **Movie 2**.

### Change point analysis to measure day of peak incidence

We used multiple change point analysis to capture the inflection points of each wave. We fit a Bayesian change point model to the log of 7-day moving average case rates for each state. The change point model consisted of seven change points and eight segments. To capture the sequential 4 wave patterns, we constrained the slopes of the four linear segments preceding each change point to be positive (ascending arm) and the slopes of the four segment following the change point to be negative (descending arm). To obtain the posterior distributions of the wave peaks, we used the package mcp in R (*4*). **Fig. S4** shows the results of the change point analysis. This figure shows 7 change points. The odd change points 1, 3, 5 and 7 represent the peak days of waves 1 through 4, respectively, and their heights represent their amplitude. The 95% credible intervals were obtained from the posterior distribution of these change points.

### Population-weighted average temperature by state

We used Daymet Version 4 Monthly Latency Daily Surface Weather Data (*5*) to obtain minimum and maximum daily temperature data on a 1 squared kilometer grid. We computed daily average temperature by taking midpoint temperature between the minimum and maximum daily temperatures. To adjust the average temperature for the populated areas at the state level, we weighted the daily average temperature by the estimated proportion of the state population living in each squared kilometer in that state (*6*). Then, we computed the average state temperature by summing these weighted temperature values across the squared kilometers in each state. Finally, we computed the 7-day moving average to smooth the temperature trends.

### Political party state government control

We examined the relationship between the political control of states’ governments and the amplitude of each wave. We used the National Conference for State Legislature (NCSL) to define the state’s government control based on the legislature’s partisan composition and the political party of the state governor. In 2020, there were 23 states controlled by the Republican party, 15 states controlled by the Democratic party, and 13 states where the state control were split. Our hypothesis was the amplitude of a wave is similar for neighboring states with similar government control and different for states with different state control. We examined 101 unique pairs of state neighbor, 51 of which had similar political controls and 60 had different controls. We defined Republican-Republican, Democratic-Democratic and Split-Split controls between a pair as similar and everything else as different. The outcome of interest was the difference in amplitude between the states in each pair. Because these differences were not normally distributed, we used the Wilcoxon signed-ranked test to test for statistical significance. We assessed whether the rank mean of the amplitude difference was statistically significantly different between states with similar government control vs those that have dissimilar government controls.

**Fig. S1.**
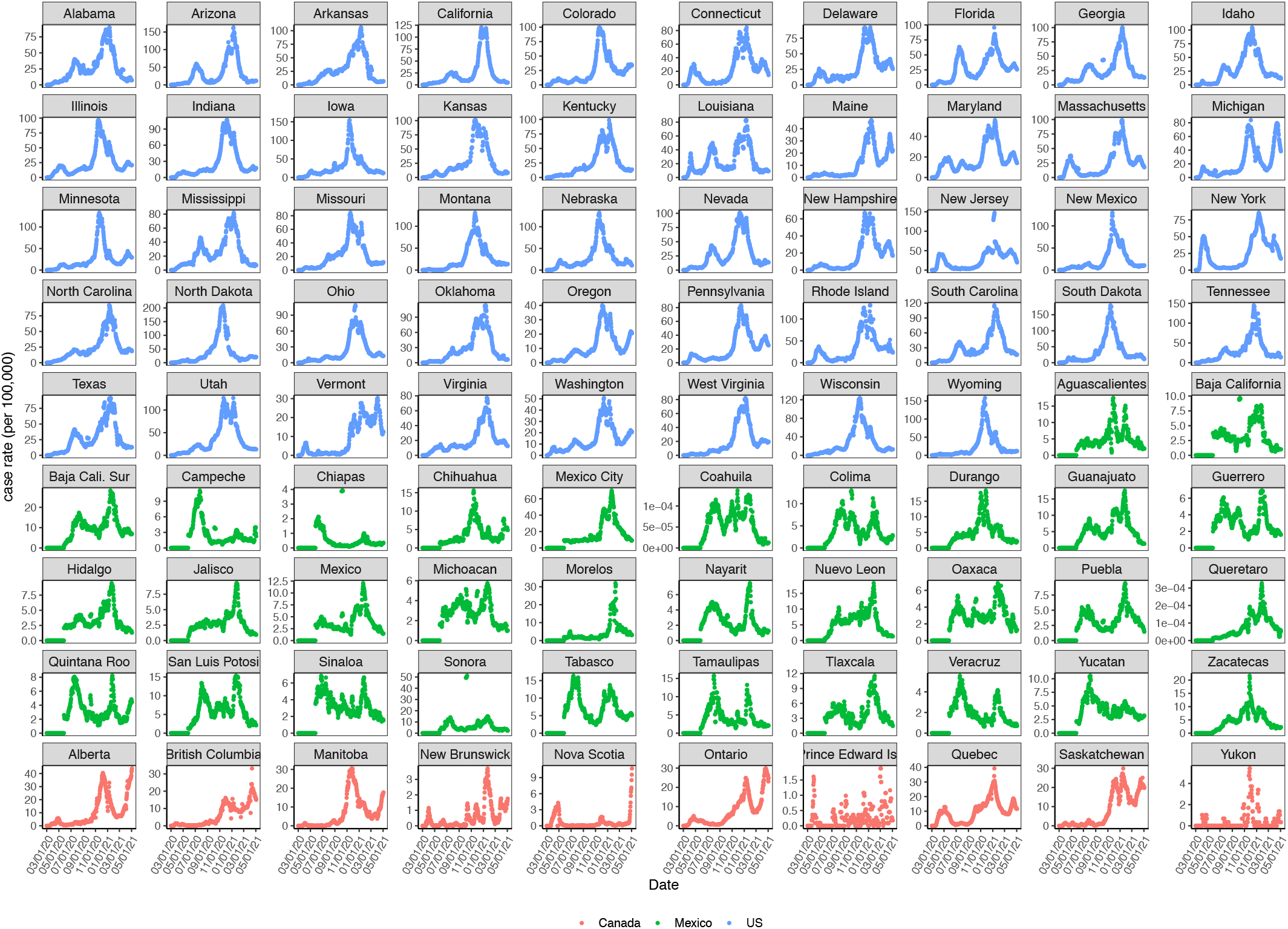
Covid-19 case incidence rates (per 100,000) by state in the US (blue), Mexico (green), and Canada (red). Incidence rates are smoothed by taking 7-day moving average.

**Fig. S2.**
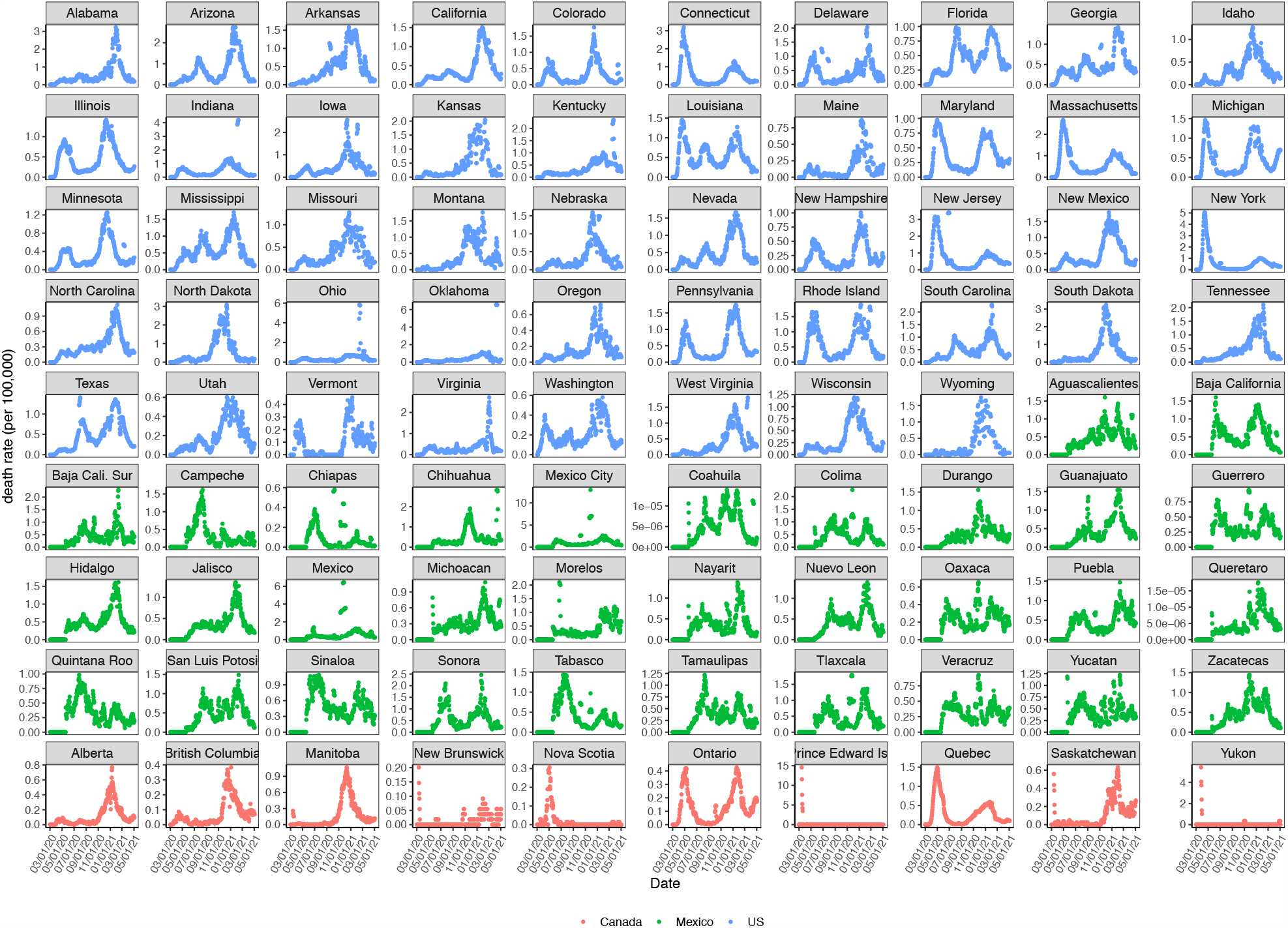
Covid-19 mortality rates (per 100,000) by state in the US (blue), Mexico (green), and Canada (red). Mortality rates are smoothed by taking 7-day moving average.

**Fig. S3.**
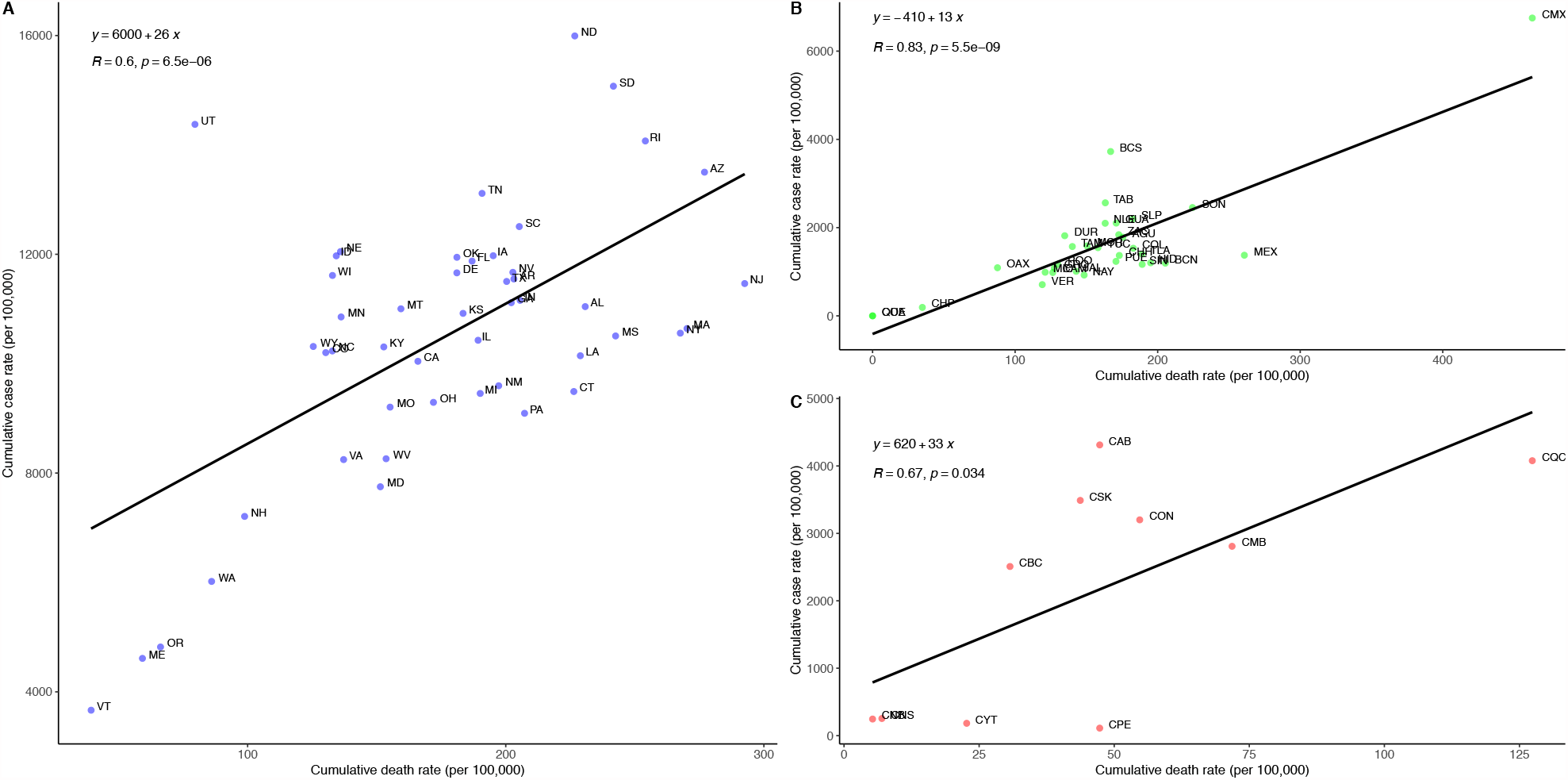
Correlation of covid-19 cumulative incidence and mortality rate in the US. Across the 48 states in the US, we observed a positive correlation (r=0.6, p < 0.01) between cumulative incidence rate and cumulative mortality rate of covid-19 from March 1,2020 to May 10, 2021. This indicates that US states with higher cumulative case rate tend to have higher cumulative mortality rates from covid-19 epidemics. The slope of the fitted linear line showed the cumulative case rates that corresponds to 1 cumulative death rate, or the ratio of total number of cases to total deaths. In the US, 26 reported incidence of covid-19 corresponds to 1 death. Utah showed the highest case-to-death ratio, compared to other states. Some northeastern states (VT, ME, NH), Pacific states (WA, OR), and midwestern states (SD, ND) showed higher ratio compared to the average. In Mexico, cumulative case rate was highly correlated with cumulative death rate (r = 0.83, p <0.01). The case to death ratio was lower than the ratio in the US, indicating higher risk of death given one covid-19 case. In the 10 states of Canada, cumulative case rate was positively correlated with cumulative mortality rate. The case to death ratio was higher than the other two countries.

**Fig. S4.**
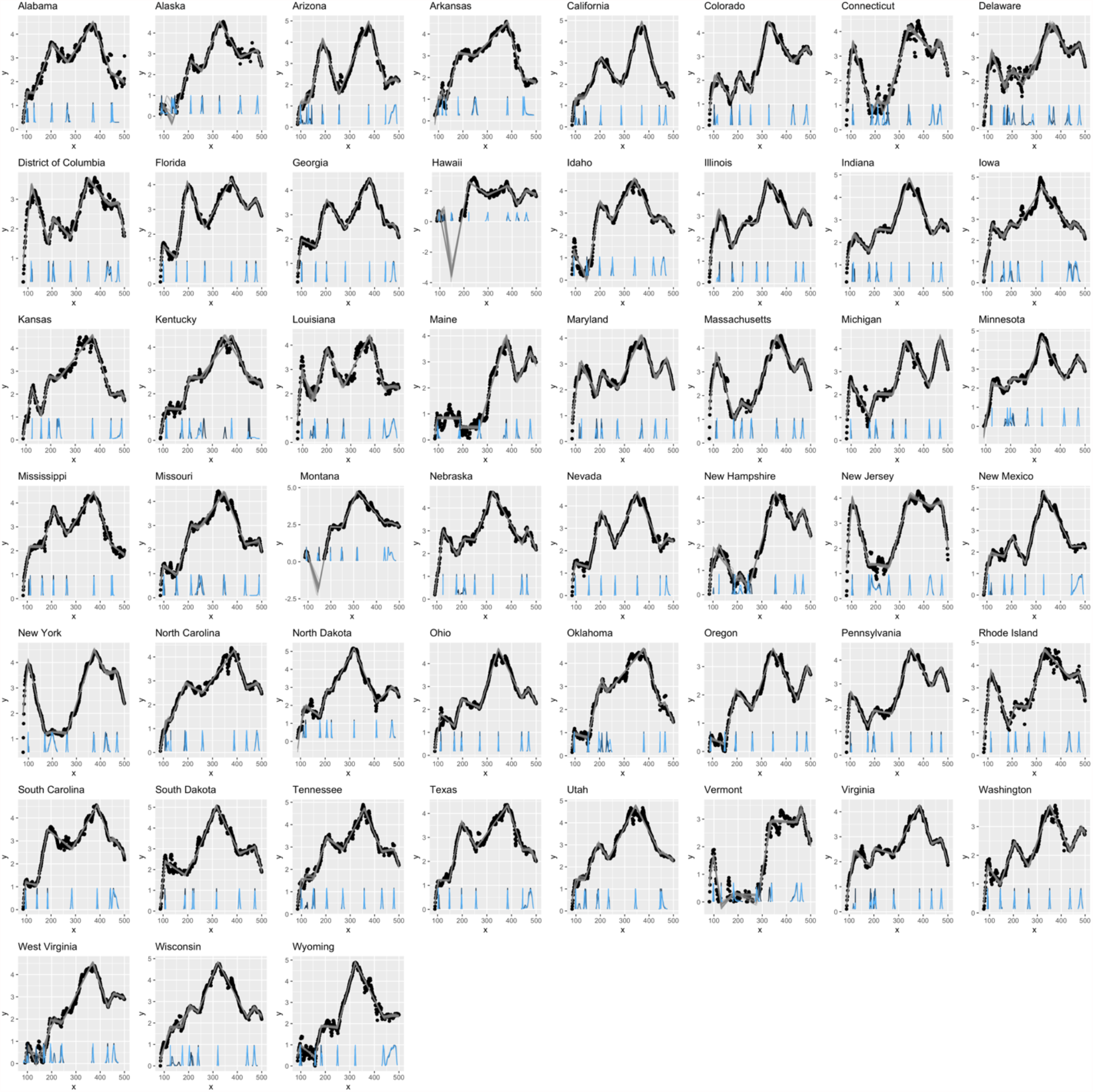
Results of the multiple change point analysis by state. The x-axis is the days since January 1^st^, 2020, and the y-axis is the log of 7-day moving average of case rates per 100,000. The gray lines are samples from the posterior fitted models, and the density plots at the bottom are the posterior densities of the seven change points. We used change points 1, 3, 5 and 7 to represent the days in which waves 1 through 4 peak, respectively.

**Fig. S5.**
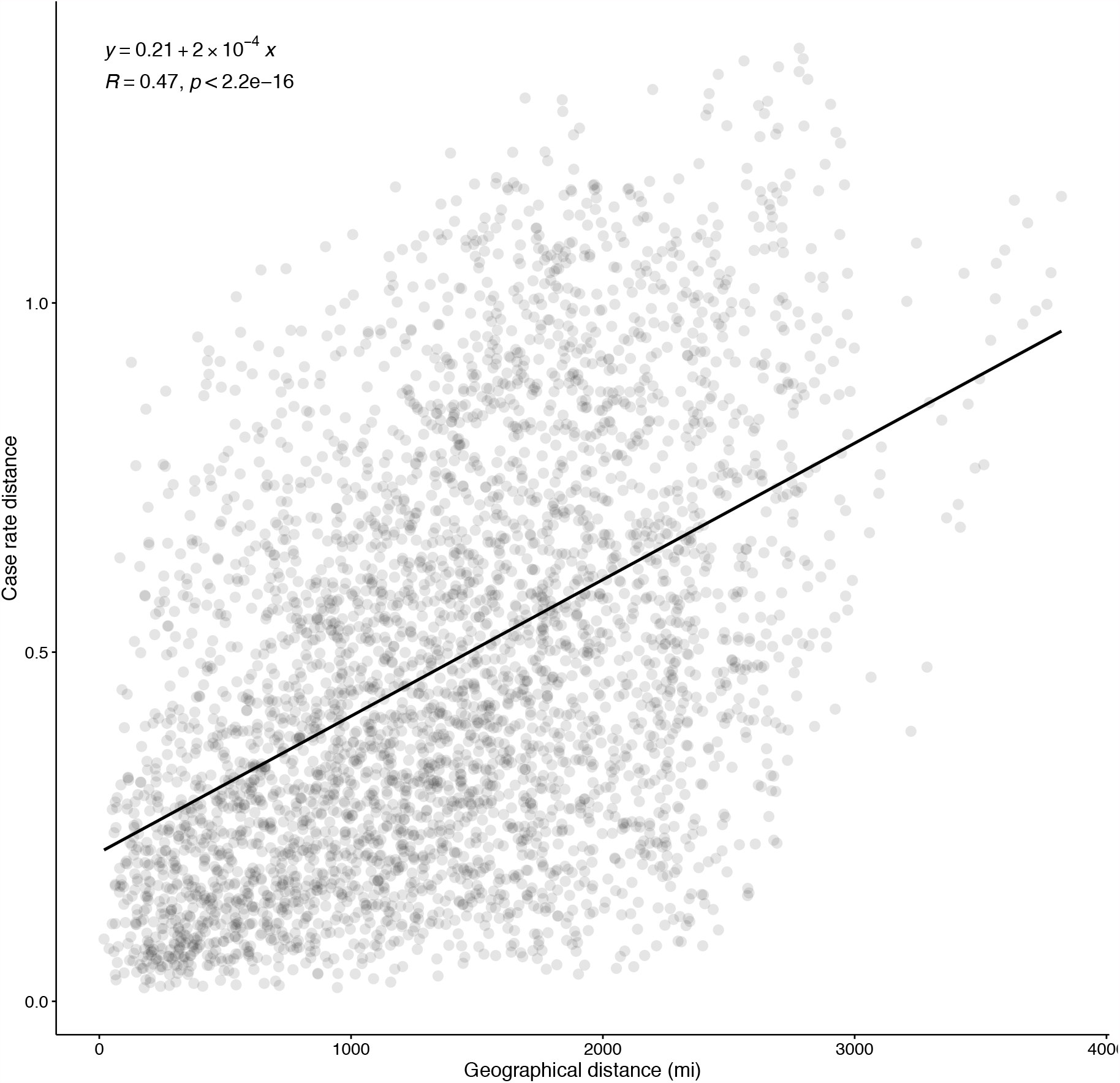
Correlation between the distance of covid-19 case rates and geographical distance for all possible pairs of US and Mexican states and Canadian provinces. The linear fit, correlation coefficient (R) and the p-value of the fit are shown.

**Fig. S6.**
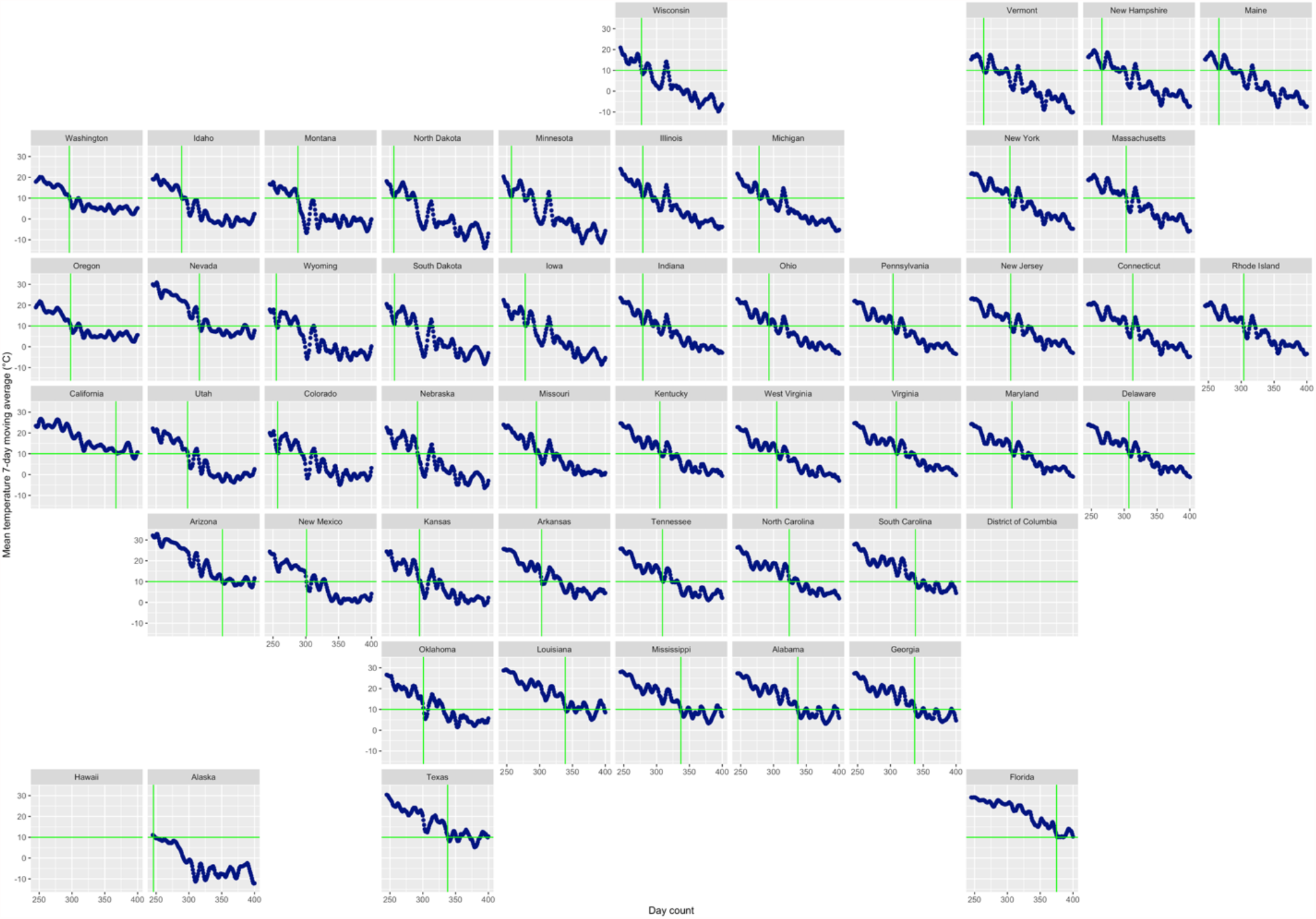
Measuring day when temperature reaches a threshold. The x-axis is day counts since January 1^st^ 2020, and the y-axis is the mean temperature 7-day moving average. Each figure represents a state which are organized roughly by their locations in the US geological map. Data for the District of Columbia and Hawaii were not available. The horizontal green line indicates the temperature threshold used in this example (10°C), and the vertical line indicates the day at which the state’s curve reaches that threshold within +/-1°C.

**Table S1.** List of state and province acronyms used in the analysis and figures.

| United States |  | Mexico |  | Canada |  |
| --- | --- | --- | --- | --- | --- |
| State | Abbr. | State | Abbr. | Province | Abbr |
| Alabama | AL | Aguascalientes | AGU | Alberta | CAB |
| Arizona | AZ | Baja California | BCN | British Columbia | CBC |
| Arkansas | AR | Baja California Sur | BCS | Manitoba | CMB |
| California | CA | Campeche | CAM | New Brunswick | CNB |
| Colorado | CO | Chiapas | CHP | Nova Scotia | CNS |
| Connecticut | CT | Chihuahua | CHH | Ontario | CON |
| Delaware | DE | Ciudad de Mexico | CMX | Prince Edward Island | CPE |
| Florida | FL | Coahuila | COA | Quebec | CQC |
| Georgia | GA | Colima | COL | Saskatchewan | CSK |
| Idaho | ID | Durango | DUR | Yukon | CYT |
| Illinois | IL | Guanajuato | GUA |  |  |
| Indiana | IN | Guerrero | GRO |  |  |
| Iowa | IA | Hidalgo | HID |  |  |
| Kansas | KS | Jalisco | JAL |  |  |
| Kentucky | KY | Mexico | MEX |  |  |
| Louisiana | LA | Michoacan | MIC |  |  |
| Maine | ME | Morelos | MOR |  |  |
| Maryland | MD | Nayarit | NAY |  |  |
| Massachusetts | MA | Nuevo Leon | NLE |  |  |
| Michigan | MI | Oaxaca | OAX |  |  |
| Minnesota | MN | Puebla | PUE |  |  |
| Mississippi | MS | Queretaro | QUE |  |  |
| Missouri | MO | Quintana Roo | ROO |  |  |
| Montana | MT | San Luis Potosi | SLP |  |  |
| Nebraska | NE | Sinaloa | SIN |  |  |
| Nevada | NV | Sonora | SON |  |  |
| New Hampshire | NH | Tabasco | TAB |  |  |
| New Jersey | NJ | Tamaulipas | TAM |  |  |
| New Mexico | NM | Tlaxcala | TLA |  |  |
| New York | NY | Veracruz | VER |  |  |
| North Carolina | NC | Yucatan | YUC |  |  |
| North Dakota | ND | Zacatecas | ZAC |  |  |
| Ohio | OH |  |  |  |  |
| Oklahoma | OK |  |  |  |  |
| Oregon | OR |  |  |  |  |
| Pennsylvania | PA |  |  |  |  |
| Rhode Island | RI |  |  |  |  |
| South Carolina | SC |  |  |  |  |
| South Dakota | SD |  |  |  |  |
| Tennessee | TN |  |  |  |  |
| Texas | TX |  |  |  |  |
| Utah | UT |  |  |  |  |
| Vermont | VT |  |  |  |  |
| Virginia | VA |  |  |  |  |
| Washington | WA |  |  |  |  |
| West Virginia | WV |  |  |  |  |
| Wisconsin | WI |  |  |  |  |
| Wyoming | WY |  |  |  |  |

## Notes

### Competing Interest Statement

The authors have declared no competing interest.

### Author Declarations

Not human subject research. All data were publicly available and de-identified. IRB was not required.

